# Covid-19 related stress and coping strategies among adults with chronic disease in Southwest Ethiopia

**DOI:** 10.1101/2020.08.14.20174318

**Authors:** Abel Girma, Ermias Ayalew

## Abstract

**Background:** Coronavirus disease (COVID-19) is an infectious disease caused by a newly discovered coronavirus which are a large family of viruses that are common in people and many different species of animals which can affect people physically and psychologically. The older people and those with underlying medical problems are more likely to develop serious illness and death.

**Objective:** the aim of this study was to determine Covid-19 related stress and coping strategies among adults with chronic disease in Bench-Sheko,West Omo and Keffa Zones, southwest Ethiopia

**Methods:** Institutional based cross-sectional study was applied among 613 adults with chronic disease. A simple sampling technique was applied. Correlational analysis was used to determine the relationship between the COVID-19 related stress score and coping strategy types. To measure the strength of association between dependent variables and independent variables and Pearson coefficients (r) with 95% Confidence interval (CI) were calculated. Finally, the variable, which shows statistical significance (p-value < 0.05 cut point) were used to quantify the associations among variables.

**Results:** Around 613 participants of 96% response rate were participated. About 68.4% were moderately stressed; low stress was 17.8% and severe stress was 13.9 %. Covid-19 related perceived stress score were positively associated with coping strategies types of like sell-distraction, active coping, denial, emotional support, behavioral disengagement, venting, and use of instrument, positive reframing, self-blaming, planning, humoring and religion. The most preferable types of coping strategies were religious, instrumental and active coping strategies and while the least used were substance used.

**Conclusion:** Significant numbers of participants were suffered from severe perceived stress due to covid-19 outbreak in this study area. Both adaptive and maladaptive Coping strategy types were significantly associated with stress. Substance use and self-blaming were the types coping strategies which were not associated with perceived stress.

## Introduction

Coronavirus disease (COVID-19) is an infectious disease caused by a newly discovered coronavirus which are a large family of viruses that are common in people and many different species of animals, like camels, cattle, cats, and bats. Sometimes, animal coronaviruses can infect people and then spread between people like with MERS-CoV, SARS-CoV, and now with SARS-CoV-2[1, 2]. The pandemic of corona virus disease have been affecting people both physically and psychologically. So that many people have been experienced stress, anxiety and depression reactions[3].

Stress during an infectious disease outbreak can include fear and worry about own health, worsening of chronic health problems, worsening of mental health condition and also increased use of alcohol and tobacco[4]. Most people infected with the COVID-19 virus will experience mild to moderate respiratory illness and recover without requiring special treatment. However, the older people and those with underlying medical problems like cardiovascular disease, diabetes, chronic respiratory disease, and cancer are more likely to develop serious illness and death[1]. The mental health and psychosocial consequences of the COVID-19 pandemic may be more serious among those who have been in contact with the virus; those who are vulnerable to biological or including people affected by mental health problems; health professionals; and even people who are following the news through numerous media channels[5].

The corona virus disease outbreak is leading to health problems like stress, anxiety, depressive symptoms, insomnia, denial, anger and fear globally. The pandemic and the containment measure like quarantine, social distancing, and self-isolation can also have a detrimental impact on mental health. The increased loneliness and reduced social interaction are risk factors for many mental disorders. The collective concerns influence daily behaviors, economy, prevention and decision making form health organization, policy makers and medical centers, which can weaken strategies of corona virus disease control and leads to more morbidity and mental health needs at global level[5, 6].

Assessment of COVID-19 related stresses and coping strategy are a paramount at several levels. The finding will be used to give appropriate intervention of COVID-19 related stresses and coping strategy, it will provide sufficient data for any concerning body like health professionals, policy makers and planers in advising the most vulnerable population for prevention and control of COVID-19 physiological and psychological impacts timely and appropriately. It will fill the information gap of the study area and it will provide a base line information for others researcher to conduct similar study by considering the limitation of this study.

Globally the battle against physiological and psychological impact of COVID-19 is still continuing. Some unprecedented measures have been adopted to control the COVID-19 transmission in all parts of Ethiopia, including the suspension of public transportation, the closing of public spaces, close management of communities, and isolation and care for infected people and suspected cases. The global mental health issues related to corona virus disease outbreak may evolve into long-lasting health problems, isolation and stigma. the global health intervention should be employed to address psychosocial stressors, particularly related to the use of isolation/quarantine, fear and vulnerability among the general population like elder and those who have chronic disease [7]. The outbreak of coronavirus disease 2019 (COVID-19) may be stressful for people but coping with stress will make stronger[4]. Different people are reacting differently to stressful situations. These situations mainly depend on the background of the person, the things that make the people different from other peoples, and the community they live in. The people who may respond more strongly to the stress of an outbreak are older people and those with chronic disease[4]. The people at higher risk for severe illness like people with chronic disease are at increased risk of stress due to corona virus disease outbreaks. Even though some studies conducted in different parts of the world, they were mainly focused on general population which lacks representation of the main risky population for both physiological and psychological risks like individuals with chronic diseases which are unable to access easily needed information and also routine health service are disrupted due to corona virus disease outbreak[8]. In addition to managing their existing health conditions, coping with the stress associated with the COVID-19 outbreak may be more challenging for people with chronic disease. This suggests that understanding the perceived impact of pandemic can inform the development of more adequate care and support, interventions, psycho education, and services for people with chronic conditions. Moreover they are no any study conducted before regarding to these risky groups in the study area. Therefore to facilitate outbreak prevention and control of COVID-19 in Ethiopia, in particulars of Bench Sheko Zone, Kaffa Zone and West Omoo zone, there was an urgent need to understand the perceived stress level and coping strategies types, in particular, of those who are most vulnerable, of COVID-19 at this most critical time because of the physiological and psychological long lasting impact of COVID-19. So that, this study was aimed to investigate COVID-19 related perceived stress level and coping strategies types among adults with chronic disease who live in the three zone of southwest Ethiopia.

## Materials and methods

### Study design

Institutional based cross-sectional study was applied.

### Sample size determination

The required sample size of the study participants was determined by population proportion formula of.

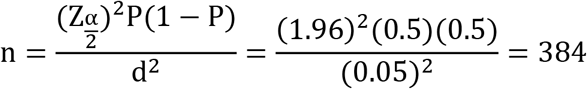

Assumptions;

n = the number of participant to be interviewed

Z = standardized normal distribution value at the 95% CI, which is 1.96

P = 50% was considered

d = the margin of error, taken as 5%

Because of the sampling procedure design effect of 1.5 was used.

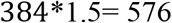

After adding, a 10% of non-response rate the final sample size became 634.

### Sampling technique

First, three district was selected from each zone by using simple random sampling technique and then the health facility from each selected district was selected again by simple random sampling technique. After that, the calculated sample size was proportionally allocated to randomly selected public health institutions in the three zones based on the number of chronic disease who attending each health facility. To select study subjects from each unit of health care service simple random sampling technique was applied by using client’s registrations number.

### Data collection and analysis

The data were collected by using structured interviewer administered questionnaire by well-trained health professionals. Socio-demographics variables like age, residence, ethnicity, religion, marital status, educational status, occupation, income, COVID-19 related stress, types of coping strategies of adults with chronic disease were assessed.

All the data were checked for completeness and internal consistency by cross checking and then was coded entered into Epi Data 4.6.0.2 computer software package and cleaned for inconsistency. For further analysis, the data were exported to Statistical Package for Social Science (SPSS) version 21 software. The descriptive analyses of data were indicated using numerical summary measures and the data were presented using frequency tables, figures and graphs. The correlational analysis was used to determine the association between the COVID-19 related stress score and coping strategy types among adults with chronic disease. To measure the strength of association the Pearson coefficients (r) with 95% Confidence interval (CI) were calculated. Finally, the variable, which shows statistical significance (p-value < 0.05 cut point) were used to quantify the associations among variables.

### Data collection instruments

The COVID-19 related stress was assessed by using the Perceive Stress Scale (PSS)) which was adapted from previous study. The perceived stress scale which consists of 10 questions was used to assess the perceived stress level. PSS is a 10 item scale where respondents rate themselves in a five point scale (i.e. 0 = Never, 1 = Almost Never, 2 = Sometimes, 3 =Fairly Often, 4 = Very Often). PSS scores were obtained by reversing responses (e.g., 0=4, 1=3, 2=2, 3=1& 4=0) to the four positively stated items (4, 5, 7 and 8). Total scores would be obtained by summing all scale items. Finally categorized as low Stress: those who scored ranging from 0-13 on stress questions, moderate Stress: those who scored ranging from 14-26 on stress questions and high perceived Stress those who scored ranging from 27-40 on stress questions[9, 10]

The coping strategies was assessed by using the Brief COPE Inventory scale.The Brief COPE Inventory consists of 28 statements, across two scales, which can identify around 14 possible coping strategies of stress but due to shortage of time we used 14 questions. It is focused mainly on understanding the frequency with which people use different coping strategies in response to various stressors. Participants using the inventory, score themselves from 1 to 4 with 1 being ‘I *haven’t been doing this at all* and 4 being ‘*I’ve been doing this a lot*. The higher scores was when the participants give the response of 3 or 4 for the given statement and the individually was considered as highly using specific coping strategy as one of his core coping strategies. If they score low - with a 1 or 2 - then we couldn’t consider as one of his core coping strategies[10, 11]. The Brief COPE Scores Scales were computed with no reversals of coding and it includes Self-distraction, Active coping, Denial, Substance use, Use of emotional support, Use of instrumental support, Behavioral disengagement, Venting, Positive reframing, Planning, Humor, Acceptance, Religion and Self-blame[10, 11].

To assure the quality of the data, pre-structured questionnaire was used. The final version of the questionnaire which was prepared in English were translated into the local language of the respondents (Amharic) and again translated back to English. Both the data collectors and facilitators were local languages speakers. The data collectors and supervisors were given one day intensive training by principal investigator (PI) on the instruments, method of data collection, ethical issues and the purpose of the study. Intensive supervision was done by principal investigator and supervisor and they were checked the collected data for completeness, accuracy, and consistency throughout the data collection period. The overall supervisions were done by the principal investigators.

## Results

### Socio-demographic characteristic

From the total of 634 sample sizes, 613 (96%) were participated in this study. The majority 379 (61.8%) were males with the mean age of 36.93 (SD: 1.677). Around 390 (63.6%) have been lived in the urban region, and 161 (26.3%) are belongs to Amhara ethnic. Regarding to their religion and marital status 272 (44.4%) are Protestant religion followers & 405 (66.1%) were married. Concerning the educational status 201 (32.8%) are able to Read & write & while 147 (24%) of the participants are Farmer by their occupation (see Table 1).

**Table 1:** shows socio-demographic characteristics of study participants in Bench-Sheko,West Omoo and Keffa zone, southwest Ethiopia(N=613).

| Variables | Frequency | Percent % |
| --- | --- | --- |
| Age |  |  |
| 18-29 | 180 | 29.4 |
| 30-49 | 332 | 54.2 |
| >50 | 101 | 16.5 |
| Sex |  |  |
| Male | 379 | 61.8 |
| Female | 234 | 38.2 |
| Residence |  |  |
| Urban | 390 | 63.6 |
| Rural | 223 | 36.4 |
| Ethnicity |  |  |
| Bench | 104 | 17.0 |
| Kaffa | 160 | 26.1 |
| Sheka | 13 | 2.1 |
| Menit | 125 | 20.4 |
| Amhara | 161 | 26.3 |
| Others | 50 | 8.2 |
| Religion |  |  |
| Orthodox | 268 | 43.7 |
| Protestant | 272 | 44.4 |
| Muslim | 63 | 10.3 |
| Catholic | 5 | .8 |
| Other | 5 | .8 |
| Marital status |  |  |
| Married | 405 | 66.1 |
| Single | 135 | 22.0 |
| Divorced | 53 | 8.6 |
| Widowed | 14 | 2.3 |
| Other | 6 | 1.0 |
| Educational status |  |  |
| Cannot read & write | 165 | 26.9 |
| Read & write only | 201 | 32.8 |
| Primary school | 95 | 15.5 |
| Secondary school | 83 | 13.5 |
| Tertiary education | 69 | 11.3 |
| Occupation |  |  |
| Student | 54 | 8.8 |
| Farmer | 147 | 24.0 |
| Merchant | 95 | 15.5 |
| Housewife | 98 | 16.0 |
| Governmental employ | 119 | 19.4 |
| Private employ | 65 | 10.6 |
| Daily laborer | 18 | 2.9 |
| Other | 17 | 2.8 |
| Income |  |  |
| <3000 | 470 | 76.7 |
| 3000-5000 | 101 | 16.5 |
| >5000 | 42 | 6.9 |
| Family size |  |  |
| 1-4 | 372 | 60.7 |
| ≥5 | 241 | 39.3 |

### Health related characteristic

All participants had at least 1 chronic condition, and only 15 (2.4 %) were living with 3 chronic conditions. The most common chronic conditions were HIV/AIDS (23.2%) and Diabetes (17.0%) respectively. The majority of the participants 580(94.6%) were non-smokers. The most commonly stated sources of knowledge were TV/radio (66.6 %), Health professionals (66.4%) (see Table 2).

**Table 2:**
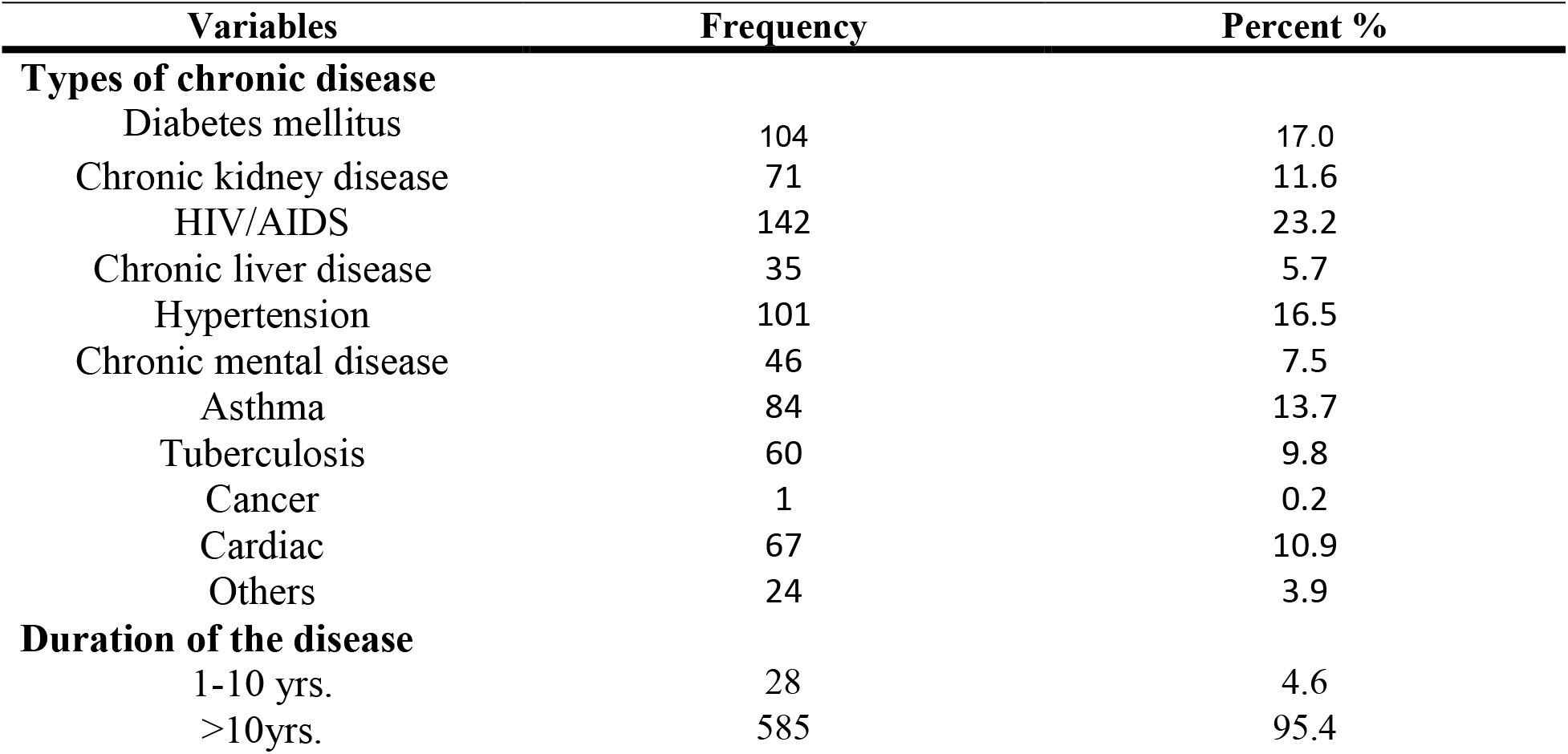
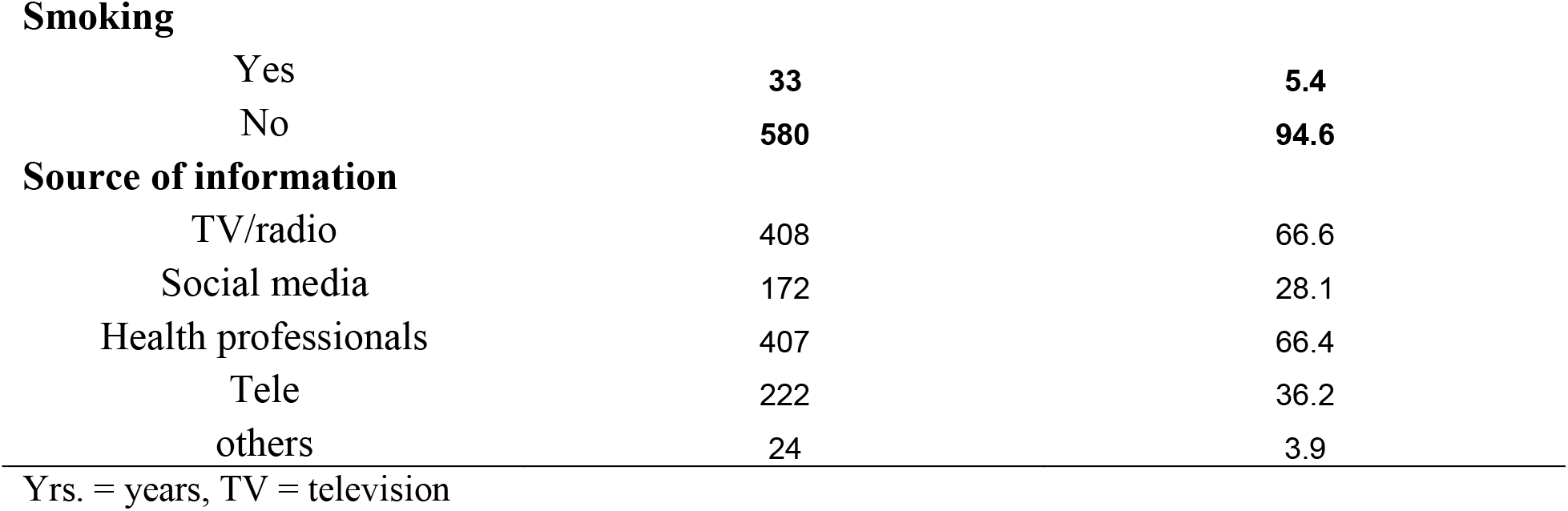
shows health related characteristic of study participants in Bench-Sheko, West Omo and Keffa zones, southwest Ethiopia

| Variables | Frequency | Percent % |
| --- | --- | --- |
| **Types of chronic disease** |  |  |
| Diabetes mellitus | 104 | 17.0 |
| Chronic kidney disease | 71 | 11.6 |
| HIV/AIDS | 142 | 23.2 |
| Chronic liver disease | 35 | 5.7 |
| Hypertension | 101 | 16.5 |
| Chronic mental disease | 46 | 7.5 |
| Asthma | 84 | 13.7 |
| Tuberculosis | 60 | 9.8 |
| Cancer | 1 | 0.2 |
| Cardiac | 67 | 10.9 |
| Others | 24 | 3.9 |
| **Duration of the disease** |  |  |
| 1-10 yrs. | 28 | 4.6 |
| >10yrs. | 585 | 95.4 |
| **Smoking** |  |  |
| Yes | 33 | 5.4 |
| No | 580 | 94.6 |
| **Source of information** |  |  |
| TV/radio | 408 | 66.6 |
| Social media | 172 | 28.1 |
| Health professionals | 407 | 66.4 |
| Tele | 222 | 36.2 |
| others | 24 | 3.9 |

Yrs. = years, TV = television

### Covid-19 related perceived stress level

The mean perceived stress score of the participants was 19.31 (SD = 7.212). Around 255 (41.6%) of the respondents were score > 20 and while 358 (58.4%) were score ≤ 20 out of 40. The majority of (68.4%) the participants were reported moderate stress level; around 17.8% were reported low stress level and while 13.9 % were severe covid-19 related perceived stress level (see Table 3).

**Table 3:** shows covid-19 related perceived stress level of the study which conducted in Bench-Sheko,West Omo and Keffa zones, southwest Ethiopia

| Covid-19 related stress level | Frequency | Percent |
| --- | --- | --- |
| Low stress score | 109 | 17.8 |
| Moderate stress score | 419 | 68.4 |
| High stress score | 85 | 13.9 |

### Coping strategies types

From the total of 613 participants, the majority 468(76.3%) of respondent were used religious coping strategies type and the second most used types were use of instrumental coping strategy around 315(51.4%); and while 314(51.2%) of the participants were used active coping strategies. The least used types of coping strategies were substance use 37(6%) (see Table 4).

**Table 4:** shows coping strategies types of the study which conducted in Bench-Sheko,West Omo and Keffa zones southwest Ethiopia

| Types of coping strategies | No. (% of )used | No. (%) of not used |
| --- | --- | --- |
| Self-distraction | 180(29.4) | 433(70.6) |
| Active coping | 314(51.2) | 299(48.8) |
| Denial coping | 182(29.7) | 431(70.3) |
| Substance use | 37(6) | 576(94) |
| Use of emotional support | 161(26.3) | 452(73.7) |
| Behavioral disengagement | 115(18.8) | 498(81.2) |
| Venting | 249(39.2) | 373(60.8) |
| Use of instrument | 315(51.4) | 298(48.6) |
| Positive reframing | 163(26.6) | 450(73.4) |
| Sell-blaming coping | 98(16) | 515(84) |
| Planning | 179(29.2) | 434(70.8) |
| Humoring | 146(23.8) | 467(76.2) |
| Acceptance | 248(40.5) | 365(59.5) |
| Religious | 468(76.3) | 145(23.7) |

### Association between COVID-19 related stress and coping strategies

The covid-19 related perceived stress was significantly associated with coping strategies types like sell-distraction r=0.135,p <0.01,active coping r=0.131,p<0.01,denial coping r=0.338,p<0.01, use of emotional support r=0.147,p<0.01, behavioral disengagement r=0.385,p<0.01,venting r=0.235,p<0.01,use of instrumental r=0.118,P<0.01,positive reframing r=0.176,p<0.01,self-blaming r=0.284, p<0.01,planning r=0.083,p<0.05,humoring r=0.113,p<05,religion r=0.241,p<0.01 (see Table 5).

**Table 5:**
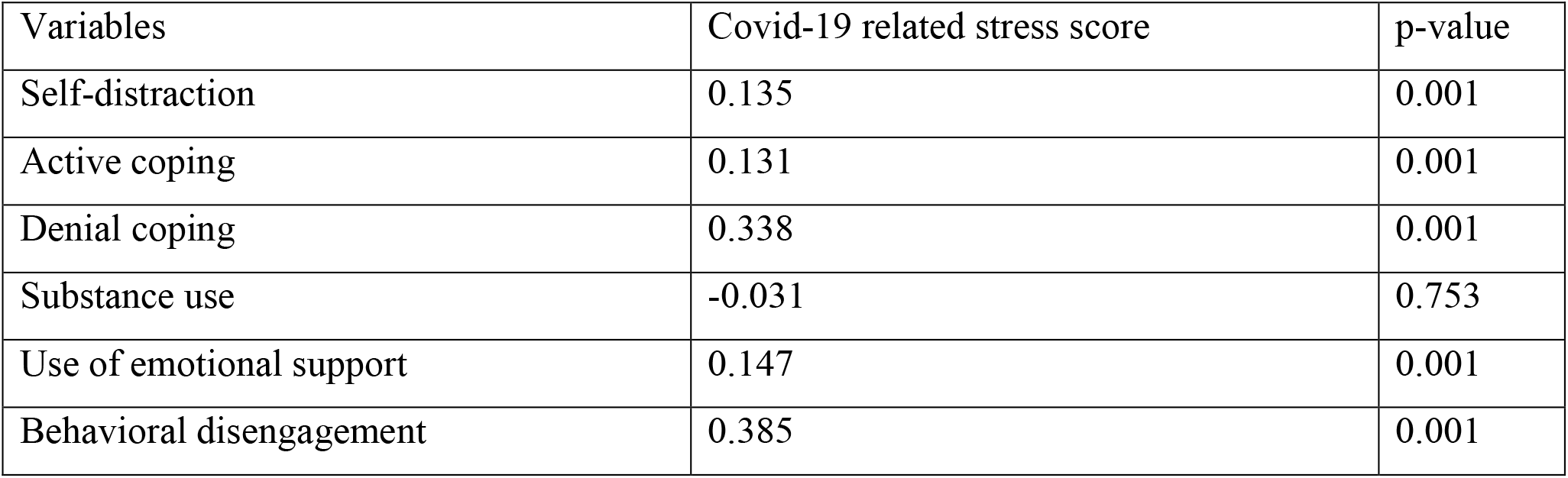
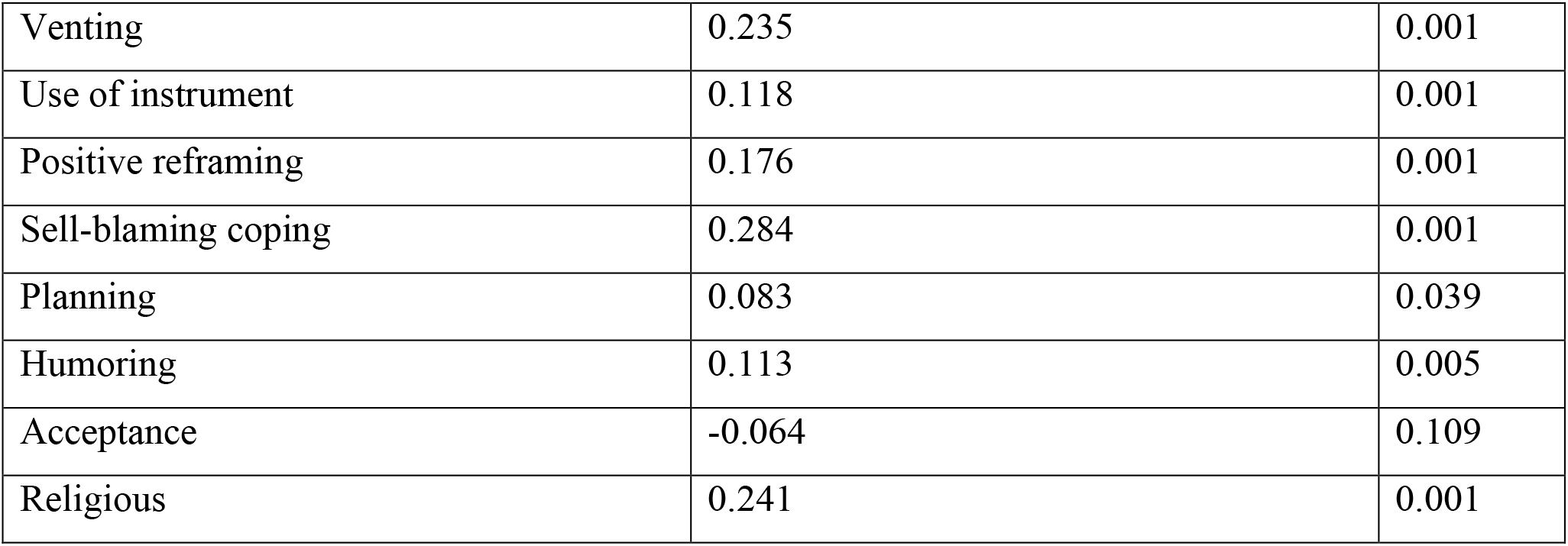
indicating the correlation between covid-19 related stress and the types of coping strategies of the study (N=613)

## Discussion

This cross-sectional study was conducted to examine the impact level of covid-19 outbreak pandemic on perceived stress and its coping strategies types which used to cope the perceived stress; and the next aim was to identify knowledge level, attitude status and practice and associated factors among chronic disease patients in Benk-Sheko, West Omoo and Kaffa Zone, southwest Ethiopia. According to this study revealed the magnitude of perceived stress were 41.6%. According to this study results showed covid-19 related perceived stress score of the participants were positively associated with coping strategies types of sell-distraction, active coping, denial, use of emotional support, behavioral disengagement, venting, use of instrument, positive reframing, self-blaming, planning, humoring and religion which mean that respondents with higher levels of covid-19 related perceived stress were used the greater levels of sell-distraction, active coping, denial coping, use of emotional support, behavioral disengagement, venting, use of instrument, positive reframing, self-blaming, planning, humoring and religion types of coping strategies to prevent and control their covid-19 related perceived stress. This finding is more likely consistent with the study conducted in unite state among individual of disability and chronic condition[10]. The participants coping strategies like substance use and acceptance were not associated with perceived stress in this study. The coping mechanism for people with disabilities and chronic illnesses may take many forms [12]. The types of coping strategies categorized in the Brief COPE as adaptive which are active coping, planning, use of emotional support, use of instrumental support, positive reframing, religion, humor, and acceptance; and maladaptive coping strategies like venting, denial, substance use, self-blame, behavioral disengagement, and self-distraction types of coping strategies[13]. According to this categorization, this study revealed that covid-19 related perceived stress score were associated with both maladaptive (i.e., denial, behavioral disengagement, venting, self-distraction, and self-blame) and adaptive (i.e., active coping, use of emotional support, use of instrument, positive reframing, planning, humoring and religion) coping strategies types among chronic disease patients. COVID-19 is an ongoing and rapidly evolving situation, which can present challenges and stressors in individuals with chronic disease. Therefore, findings from this study may help clinicians, researchers, and policymakers gain a better understanding of the use of coping strategies in individuals with chronic disease in facing this pandemic outbreak. According to this study result showed the most frequently preferable coping strategies types of the study participants were using religious, use of instrumental and using of active coping strategies for management of covid-19 related stress. Around 76.3% of the participants were used religious types of coping strategies and the second most used types of coping were use of instrumental about 51.4% and while 51.2% were used active coping strategy for managements of covid-19 related perceived stress during this pandemic outbreak. This showing that participants of this study had a tendency to cope with COVID-19 by using of religious, using of instrumental methods and involving themselves with active coping strategy. This finding is inconsistent with study which conducted in united states of America among disabled and chronic condition individual who were most frequently used acceptance and self-distracting methods to cope with covid-19 related perceived stress[14]. According to this study substance use type of coping strategy was the most least frequently used types of coping among chronic disease patient. This indicated that participants with chronic diseases were not very likely to use substance to cope with COVID-19. This finding is similar with the previous study which conducted among chronic condition and disability were used very unlikely of substance use and denial types of coping[10] and it also consistent with world health organization recommendation and center of disease control for coping of stress during covid-19 outbreak [4]

The main limitations of this study were shortage of time period for data collection which occurred due the data need urgency and the others limitation was lack of validated and reliable tools for perceived stress and coping strategies for this vulnerable groups. However some data quality control mechanisms were conducted to minimize this limitation effects on the study. Lack of previous studies also one of the main limitations. The main significance of assessment of COVID-19 related perceived stresses and coping strategy types are a paramount at several levels. One of it could be to give appropriate intervention types for COVID-19 related perceived stresses and coping strategy types of the patients about covid-19. In addition, it could provide sufficient data for any concerning body like health professionals, psychologist, policy makers and planers in advising the most vulnerable population for prevention and control of COVID-19 physiological and psychological impacts timely and appropriately. It could fill the information gap of the study area and it would be provide a base line information for others researcher to conduct similar study by considering the limitation of this study

## Conclusion

Significant numbers of participants were suffered from severe perceived stress due to covid-19 outbreak in this study area. Coping strategy types like sell-distraction, active coping, denial coping, use of emotional support, behavioral disengagement, venting, use of instrument, positive reframing, self-blaming, planning, humoring, religion were positively associated with covid-19 related perceived stress among individuals with chronic disease in this study area. Substance use and self-blaming were the types coping strategies which were not associated with perceived stress.

The most frequently used coping strategies types of the study participants were religious, using of instrumental and active coping strategies for management of covid-19 related perceived stress among individual with chronic disease while the most least used types of coping strategies were substance use.

## Data Availability

The datasets used and or analyzed during the current study are available from the corresponding author on reasonable request.

**List of abbreviations**
COVID-19: corona virus disease of 2019
SARS-COV: severe acute respiratory Syndrome
MERS-COV: Middle East respiratory syndrome
WHO: world health organization
CI: Confidence interval
CFR: case fatality ratios
SNNPR: Southern Nations, Nationalities and Peoples’ Regional State

## Ethical approval and consent to participate

After the study was approved by research directorate committee of Mizan- Tepi University, the formal letter was send for each zonal health bureau management for permission and then the informed volunteer consent was obtained from all participants.

## Consent for publication

Not applicable

## Author’s contributions

^1^was participated in data analysis, interpretation and manuscript preparation and ^2^was participated in data analysis of the study.

## Author’s information

^1^ Department of Public Health, College of Health Sciences, Mizan Tepi University, Mizan-Aman town, South Nation Nationality and People regional state, Ethiopia

^2^ Department of Nursing, College of Health Sciences, Mizan- Tepi University, Mizan-Aman town, South Nation Nationality and People regional state, Ethiopia

